# Subtracting the background: Reducing cell-free DNA’s confounding effects on *Mycobacterium tuberculosis* quantitation and the sputum microbiome

**DOI:** 10.1101/2024.03.27.24304911

**Authors:** Charissa C. Naidoo, Rouxjeane Venter, Francesc Codony, Gemma Agustí, Natasha Kitchin, Selisha Naidoo, Hilary Monaco, Hridesh Mishra, Yonghua Li, Jose C. Clemente, Robin M. Warren, Leopoldo N. Segal, Grant Theron

## Abstract

Characterising DNA in specimens from people with tuberculosis (TB), a major cause of death, is critical for evaluating diagnostics and the microbiome, yet extracellular DNA, more frequent in people on chemotherapy, confounds results. We evaluated whether nucleic acid dyes [propidium monoazide (PMA), PEMAX] and DNaseI could reduce this. PCR [16S *Mycobacterium tuberculosis* complex (*Mtb*) qPCR, Xpert MTB/RIF] was done on dilution series of untreated and treated (PMA, PEMAX, DNaseI) *Mtb*. Separately, 16S rRNA gene qPCR and sequencing were done on untreated and treated patients’ sputa before (Cohort A: 11 TB-negatives, 9 TB-positives; Cohort B: 19 TB-positives, PEMAX only) and after 24 weeks of chemotherapy (only Cohort B). PMA and PEMAX reduced PCR-detected *Mtb* DNA for both the dilution series and Cohort A sputum versus untreated controls, suggesting non-intact *Mtb* is present before treatment start. PEMAX enabled sequencing *Mycobacterium*-detection in 5/9 (59%; Cohort A) and 2/3 (67%; Cohort B week 0) TB-positive sputa where no reads otherwise occurred. In Cohort A, PMA- and PEMAX-treated versus untreated sputa had decreased α- and increased β-diversities. In Cohort B, α-diversity differences between untreated and PEMAX-treated sputa occurred only at 24-weeks and β-diversity differences between timepoints were only detected after PEMAX. DNaseI had negligible effects. PMA and PEMAX (but not DNaseI) reduced extracellular DNA, improving the proportion of *Mycobacterium* reads and PCR detection. PEMAX detected chemotherapy-associated changes in the microbiome otherwise missed. Our findings suggest these dyes improve characterization of the microbiome, especially chemotherapy-associated changes, and should be included in respiratory microbiome research in TB.

## Introduction

Tuberculosis (TB) kills an estimated 1.2 million people annually.^1^ Although sensitive tests like Xpert MTB/RIF (Xpert) are available for *Mycobacterium tuberculosis* complex (*Mtb*) detection, PCR can be confounded by DNA from non-intact cells, including from previous TB.^2–4^ This problem can be significant: in some settings up to 35% of notifications have recurrent TB; requiring special consideration in diagnostic algorithms^3^. The ability to distinguish *Mtb* DNA from old versus new episodes is therefore of clinical and epidemiological significance.

The microbiome can, in addition to the pathogen itself, influence the host immune system^7^. Characterising the role of microbial populations could unveil new targets for controlling TB. Unlike most other infectious diseases with short chemotherapy durations, TB requires ≥6 months of antibiotics with broad-spectrum rifamycins and potentially fluoroquinolones. These antibiotics have bactericidal effects,^10,11^ resulting in extracellular DNA accumulation that, in the case of *Mtb* DNA, can persist for years especially in people with compromised immunological clearance such as people living with HIV.^12^ Characterisations of the microbiome in people receiving (or who have received) TB chemotherapy thus likely measure total bacterial DNA, rather than only the subset within intact (and hence live) cells.

Viability dyes such as ethidium monoazide (EMA), propidium monoazide (PMA), and PEMAX (a EMA and PMA formulation) have been widely used for discriminating intact from non-intact microbial cells^13–17^ and act by passing through compromised membranes of non-intact cells (e.g., following heat exposure or repeated freeze-thaw^20^), after which they covalently cross-link with DNA to irreversibly form complexes not amplifiable by PCR^21^. As a result, only DNA from intact cells is, in theory, amplified once unbound dye is removed. PMA is more effective than EMA in excluding DNA from non-intact cells due to higher charge^22^. PEMAX was developed to overcome limitations of PMA and EMA such as the inability to bind DNA from cells that are still intact but no longer metabolically active and thus may improve specificity.^23^

These dyes have some data when applied to *Mtb*^24–28^. For example, the addition of PMA to Xpert shows potential for treatment monitoring, however, specificity remains an issue. More data are needed as dye penetration through *Mtb*’s waxy cell wall may be suboptimal (especially in specimens with high bacillary load)^29^ and sputum from diseased individuals contains high levels of host and microbial extracellular DNA that sequesters dye away, leaving insufficient dye to subtract background DNA. Combining viability dyes with 16S rRNA gene sequencing could improve the accuracy of microbial taxonomy by reducing background from non-intact taxa that, by virtue of their lack of viability, are unlikely to play a direct biological role.

We investigated the effects of nucleic acid dyes (PMA, PEMAX) and DNaseI on 1) *Mtb* PCR detection (16S, Xpert) in a dilution series (with versus without antibiotics, heat kill, freeze-thaw) and sputum from people with presumptive TB as well as 2) changes in the microbiome (compared by TB status and duration of chemotherapy exposure) by comparing sputum treated or untreated with dyes and DNaseI. Our overarching goal was to assess whether usage of dyes and/or DNaseI could enhance the detection of intact *Mtb* (*in vitro* and in sputum) and enhance analysis of the sputum microbiome.

## Methods

### Ethics

This study was approved by the Stellenbosch University Health Research Ethics Committee (N14/10/136, N16/05/070, M15/10/041).

### In vitro dilution series using Mycobacterium tuberculosis cells

#### Culture

*Mtb* H37Rv was cultured (37°C) to an OD_600nm_ of 1.0 in 20mL Middlebrook 7H9 liquid medium with 10% oleic acid, albumin, dextrose, and catalase (OADC; BD Diagnostics, Johannesburg, South Africa), 0.5% glycerol and 0.05% Tween-80 (Sigma-Aldrich, Modderfontein, South Africa). One millilitre aliquots were frozen (−80°C).

#### Antibiotic exposure (Experiment A)

Two hundred microlitres of thawed stock was inoculated into each of six flasks with 20mL 7H9 and incubated until mid-log (OD_600nm_ 0.6-0.8). Isoniazid (0.2g/mL in nuclease-free water; Thermo Fisher Scientific, Waltham, USA) and rifampicin (1g/mL in dimethyl sulfoxide; Sigma-Aldrich, Modderfontein, South Africa) were added to three of the 20mL culture flasks (three additional flasks served as non-antibiotic controls) (**Figure 1A**). All six flasks were incubated for 24-, 48-, and 72h at 37°C. To generate a 1:1 mixture of intact and non-intact cells, 5mL of the non-antibiotic and antibiotic-containing cultures after different incubation periods (24-, 48-, 72h) were combined in a separate sterile flask and, from each, a 100-fold serial dilution was done (10^8^, 10^6^, 10^4^ CFU/ml) in 7H9 to a total volume of 10 ml. This was done for three biological replicates.

**Figure 1.**
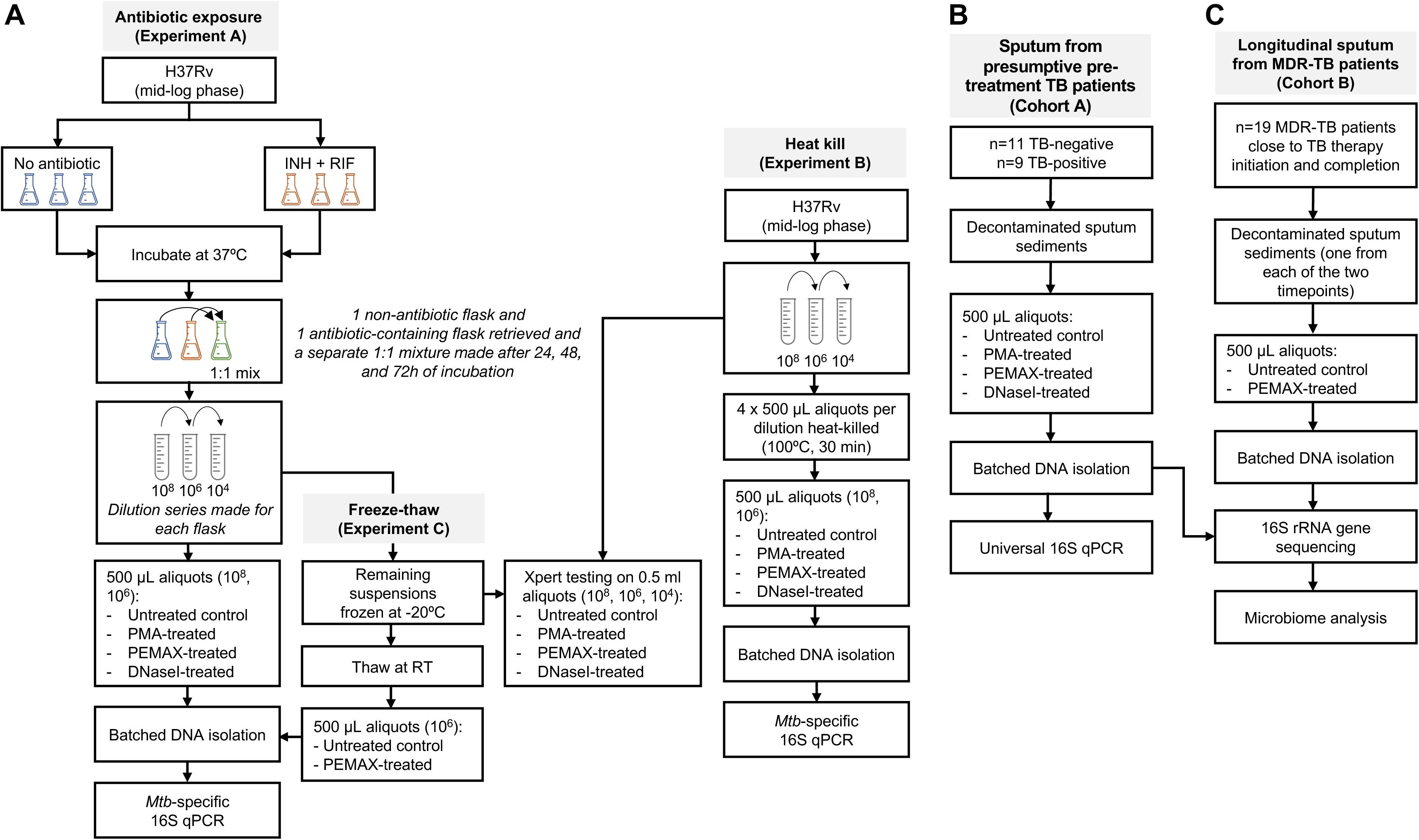
Experimental workflow for in vitro dilution series and sputum. *Mycobacterium tuberculosis* was cultured in flasks and a dilution series prepared for testing in three separate experiments: antibiotic exposure (Experiment A), heat-kill (Experiment B) and freeze-thaw (Experiment C). Sputum was collected from two distinct cohorts: one comprising patients with presumptive tuberculosis (Cohort A) and the other involving patients treated for drug-resistant tuberculosis (Cohort B). MTBC: *Mycobacterium tuberculosis* complex.

#### Heat kill (Experiment B)

For heat-kill experiments, a single culture was grown to mid-log, 100-fold dilutions (10^8^, 10^6^ CFU/ml) done and 500µL of each dilution done as stated above, heated (100°C, 30 min) and cooled to room temperature (RT). This was done for three biological replicates.

#### Freeze-thaw (Experiment C)

A freeze-thaw (FT) experiment was done with 10^6^ dilutions after 24-, 48- and 72h incubation periods that were frozen (−20°C) and allowed to thaw at RT. One biological replicate was done due to consumable unavailability.

### Treatment of samples to remove background DNA

#### PMA and PEMAX treatment (Experiments A-C)

For Experiments A and B, 500µL of each dilution was aliquoted into 1) an empty DNA LoBind tube (Eppendorf, Hamburg, Germany) without further treatment (hereafter referred to as untreated), 2) a LoBind tube containing 50µM PMA (Biotium, Fremont, USA) and 3) a PEMAX monodose tube (GenIUL, Barcelona, Spain). PMA- and PEMAX-containing tubes were vortexed for 5s before incubation in the dark (30min, 37°C). To crosslink dyes with DNA, photoactivation using the Phast Blue system (GenIUL; 60W, 2min for PMA, 15min for PEMAX) was done. PEMAX-treated samples were transferred to new LoBind tubes (PEMAX monodose tubes are hydrophilic and may cause DNA to not be neutralised by the dye^13^). The same was done for Experiment C except only PEMAX was used. At the end of dye treatment, samples were frozen (−20°C) for batched DNA isolation.

#### DNaseI treatment (Experiments A and B)

This was done for the 48h timepoint only. 500µL of each dilution was aliquoted into an empty LoBind tube (hereafter referred to as untreated) or a LoBind tube containing 1μL of RQ1 RNase-Free DNase 10X Reaction Buffer (400mM Tris-HCl, 100mM MgSO_4_, 10mM CaCl_2_;

Promega, Madison, USA) and 1μL RQ1 RNase-Free DNaseI (1U/μg DNA; Promega) and incubated (37°C, 30min) (hereafter referred to as treated). The treated tube was further incubated (65°C, 10min) with 1μL of RQ1 DNase Stop Solution and cooled to RT. Samples were frozen (−20°C) for batched DNA isolation.

### Sputum processing

#### Cohort A

Sputa from 20 people with presumptive TB were decontaminated with a modified decontamination protocol{Nikolayevskyy, 2015 #161} omitting sodium-hydroxide (to preserve bacterial viability) and an equal volume of *N*-acetyl-L-cysteine (NALC; Sigma-Aldrich) solution (0.5% w/v NALC, 2.7% w/v trisodium citrate) for 15min at RT, followed by neutralisation with a double phosphate buffer volume (BD Diagnostics) and centrifugated (3000xg, 15min). Pellets were resuspended in 5mL phosphate buffer and biobanked (−20°C) for PMA, PEMAX or DNaseI treatment as described earlier (**Figure 1B**). People classified as TB-negative (n=11) if they were MTBC culture-negative (or, if their culture was contaminated, sputum Xpert negative; n=1) or TB-positive (n=9) if they were MTBC culture-positive (or, if culture-contaminated, sputum Xpert-positive; n=2).

#### Cohort B

Sputum from 23 people with multidrug resistant (MDR)-TB at two timepoints (before or shortly after chemotherapy start, after 24 weeks chemotherapy) were decontaminated and pellets resuspended in 2mL phosphate buffer, from which one 500µL aliquot was immediately PEMAX-treated as described above and the other left as is. Both aliquots were stored at −20°C for batched DNA isolation (**Figure 1C**).

### Microbial DNA isolation

DNA was isolated from the Experiments A-C dilution series using the QIAgen DNA Mini Kit (QIAGEN, Hilden, Germany) with modifications. Briefly, aliquots were heat-killed (95°C, 15min), resuspended in lysozyme-containing (10mg/ml) lysis buffer, incubated (1h, 37°C), and proteinase K digestion done (56°C, 30min) before isolation. DNA was isolated from Cohort A and B sputum using the Purelink Microbiome DNA Purification Kit (Invitrogen, Carlsbad, USA) per manufacturer’s instructions^30^.

### 16S rRNA gene characterisation

#### Quantitative real-time PCR (qPCR) (Experiments A-C)

Reactions comprised of 5μL iTaq Universal SYBR Green (Thermo Fisher Scientific, Massachusetts, United States), 0.3μL of each forward and reverse primer (10μM), 1.4μL nuclease-free water and 3μL template DNA were done using the CFX Connect Real-Time PCR Detection System (Bio-Rad, Fremont, USA). *Mtb*-specific primers were used for the dilution series and universal 16S rRNA primers^31^ used for sputum (Cohorts A and B) (**Supplementary Table 1**). Cycling conditions were 95°C for 5min followed by 35 cycles of 95°C for 5s and 60°C for 30s. All reactions were in three technical replicates and cycle thresholds (C_T_; number of cycles before product detected) recorded. In each experiment, non-template control (NTC) C_T_s were averaged to derive a positivity threshold.

### Detection of viable bacilli by Xpert MTB/RIF (Experiments A and B)

500μL of each dilution (10^4^, 10^6^, 10^8^ CFU/ml) were Xpert-tested (**Figure 1**). Aliquots were either first treated with PMA, PEMAX or DNaseI as described above or were untreated. The manufacturer-recommended input volume of the (un)treated dilution (500µl) was used for PEMAX Xpert testing^32^. Therefore, Xpert Sample Reagent (1.5mL) was added to each 500µl aliquot and incubated (RT, 15min) with intermittent shaking before 2mL of the mixture tested (version G4; Cepheid, USA). Xpert only reports C_Tmin_ for positive results.

#### Sequencing and analysis

Sequences (V4 region, 150bp paired-end) generated by Illumina MiSeq as described ^33,34^ were demultiplexed (QIIME 2; v2020.2)^35^ and denoised (DADA2)^36^. Taxonomic classification was done with the Naïve Bayes classifier trained on Greengenes 13.8 reference database^37^ (pre-clustered at 99% identity with 515F/806R primers selected^38^). BIOM tables were rarefied by random subsampling to equal depth. Potential contaminants (in blank dyes, DNaseI and reagents) were identified using *decontam* (v1.22.0)^39^ with a threshold of 0.5 based on their prevalence in sputum vs. background samples. Contaminant amplicon sequencing variants (ASVs) were not removed but highlighted if later identified as discriminatory. α- and β-diversities were calculated using *vegan* (v2.6-4)^40^. Data and code will be available upon publication.

### Statistical analysis

The Kruskal-Wallis test with Dunn’s multiple comparisons was used to compared untreated and dye-treated C_T_ values (Experiments A-C). The Wilcoxon matched-pairs signed rank test was used for sputum qPCR (Cohort A) and Xpert results. One-sided tests were used because we hypothesised that dyes would improve *Mtb* detection. P-values are for comparisons between untreated and treated (PMA, PEMAX, or DNaseI) and were not calculated for C_T_s exceeding that of the non-template control. The Friedman test with Dunn’s multiple comparisons was used to compare α-diversity between untreated and treated sputa (Cohorts A and B). Bray-Curtis distances were compared using Kruskal-Wallis with Dunn’s multiple comparisons. *DESeq2* (v1.42.0)^41^ (with Benjamini–Hochberg multiple comparisons) was used for differential abundance testing. Feature tables were pruned to >1% relative abundance in >1% of samples. Analyses were done using GraphPad Prism (v8.0.1) and R (v4.3.2). p≤0.05 and Q-values <0.2 were significant.

## Results

### PMA and PEMAX treatment reduce *Mtb* DNA detection in dilution series (Experiments A and B)

#### 16S rRNA qPCR readouts

Dye-treated aliquots had lower detected mycobacterial load (increased C_T_) than untreated controls (**Figure 2A-B**). The same finding occurred using combinations of antibiotic-exposed or heat-killed cells, as well as different incubation periods (48h, 72h; **Supplementary Figure 1**). FT resulted in lower detected mycobacterial load [24h non-antibiotic: ΔC_T_ 2.13 (95% CI: 1.25–2.99)] compared to pre-FT samples, but when the same FT vs. pre-FT comparison was done for PEMAX-treated aliquots, a larger decrease in detected load occurred [24h non-antibiotic: ΔC_T_ 6.44 (5.53–7.36)] (**Supplementary Figure 2**).

**Figure 2.**
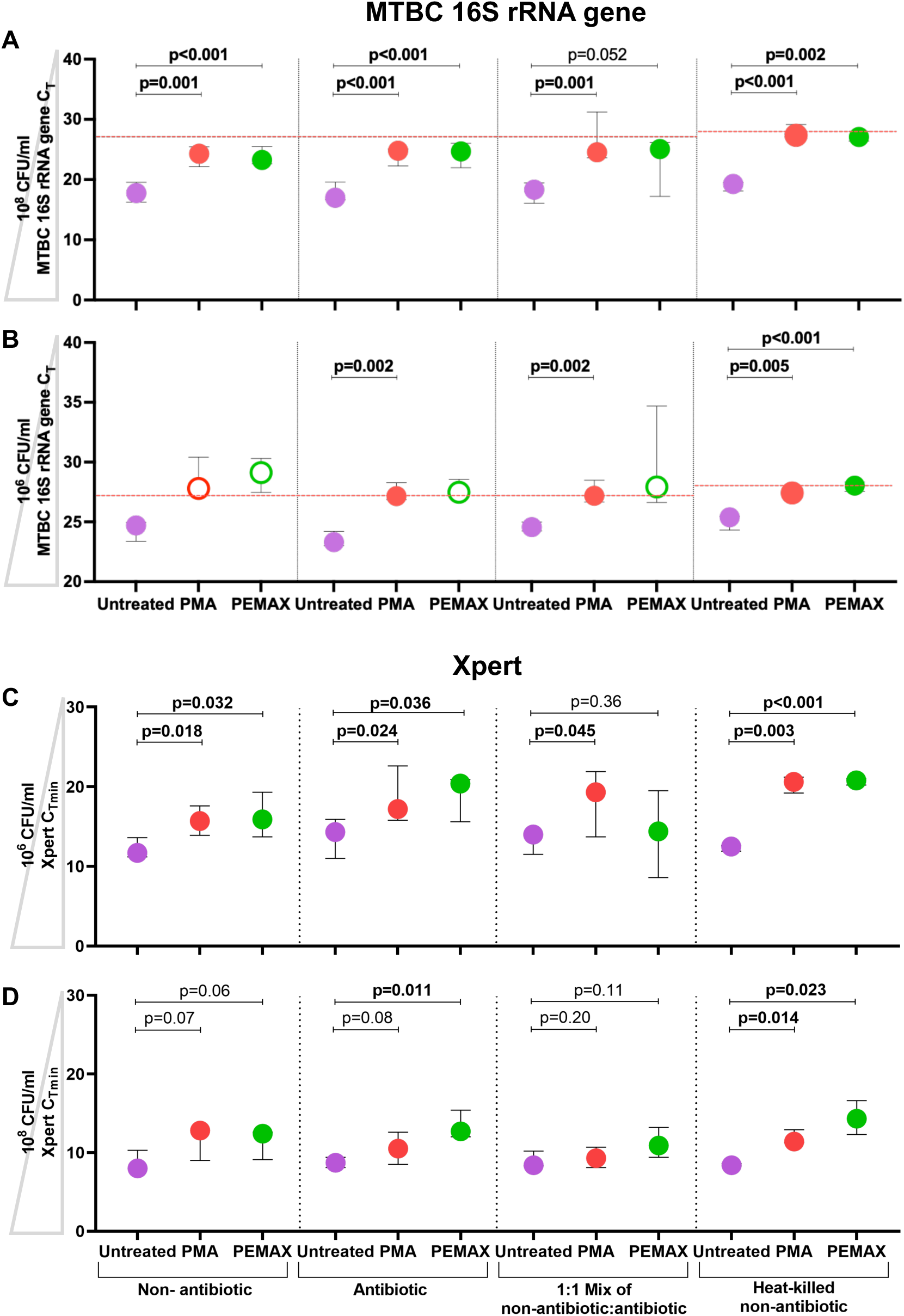
PMA and PEMAX reduce MTBC detection in dilution series (regardless of how aliquots were treated) and in sputum sediments from presumptive TB patients. PCR readouts for non-antibiotic (cultured in absence of antibiotics), antibiotic (cultured with isoniazid and rifampicin), 1:1 mixture (non-antibiotic: antibiotic), and heat-killed (non-antibiotic aliquots heated at 100°C for 30min) cultures after 24h of incubation. **Supplementary Figure 1** has these data for the 48h and 72h exposure period. Median C_T_s (IQR) are shown for **(A)** 10^8^ and **(B)** 10^6^ CFU/ml dilutions. Open circles denote C_T_s that exceed those of the averaged non-template control (dashed line). Median Xpert C_Tmin_ (95% CI) are shown for **(C)** 10^8^ CFU/ml and **(D)** 10^6^ CFU/ml. Triangles on y-axes indicate decreasing load. MTBC: *Mycobacterium tuberculosis* complex; C_T:_ cycle threshold; C_Tmin_: minimum cycle threshold;. CI: confidence interval; IQR: interquartile range.

#### Xpert

Dye reduced load readouts vs. untreated controls regardless of if aliquots underwent antibiotic or heat or no exposure (**Figure 2C-D**).

### DNaseI treatment does not reduce *Mtb* DNA detection in dilution series (Experiment A and B)

DNaseI had no effect on C_T_ compared to untreated controls. Only the antibiotic-exposed cells showed, after DNaseI treatment, unexpectedly higher detected mycobacterial load [for example, the 10^8^ CFU/ml dilution had median (IQR) C_T_s of 19.24 (19.02-19.42) vs. 19.32 (19.24-20.96) for untreated; p=0.014] (**Supplementary Figure 3**). There was no significant difference in DNaseI-exposed aliquots tested with Xpert (**Supplementary Figure 4**).

### Sputa from presumptive tuberculosis patients are abundant in extracellular bacterial DNA (Cohort A)

#### PMA, PEMAX, and DNaseI’s effect on PCR quantitation and sputum α- and β-diversity

*qPCR:* Decontaminated sputum treated with PMA or PEMAX had lower measured load than untreated controls (**Figure 3A**) whereas no differences in load resulted from DNaseI-treatment.

**Figure 3.**
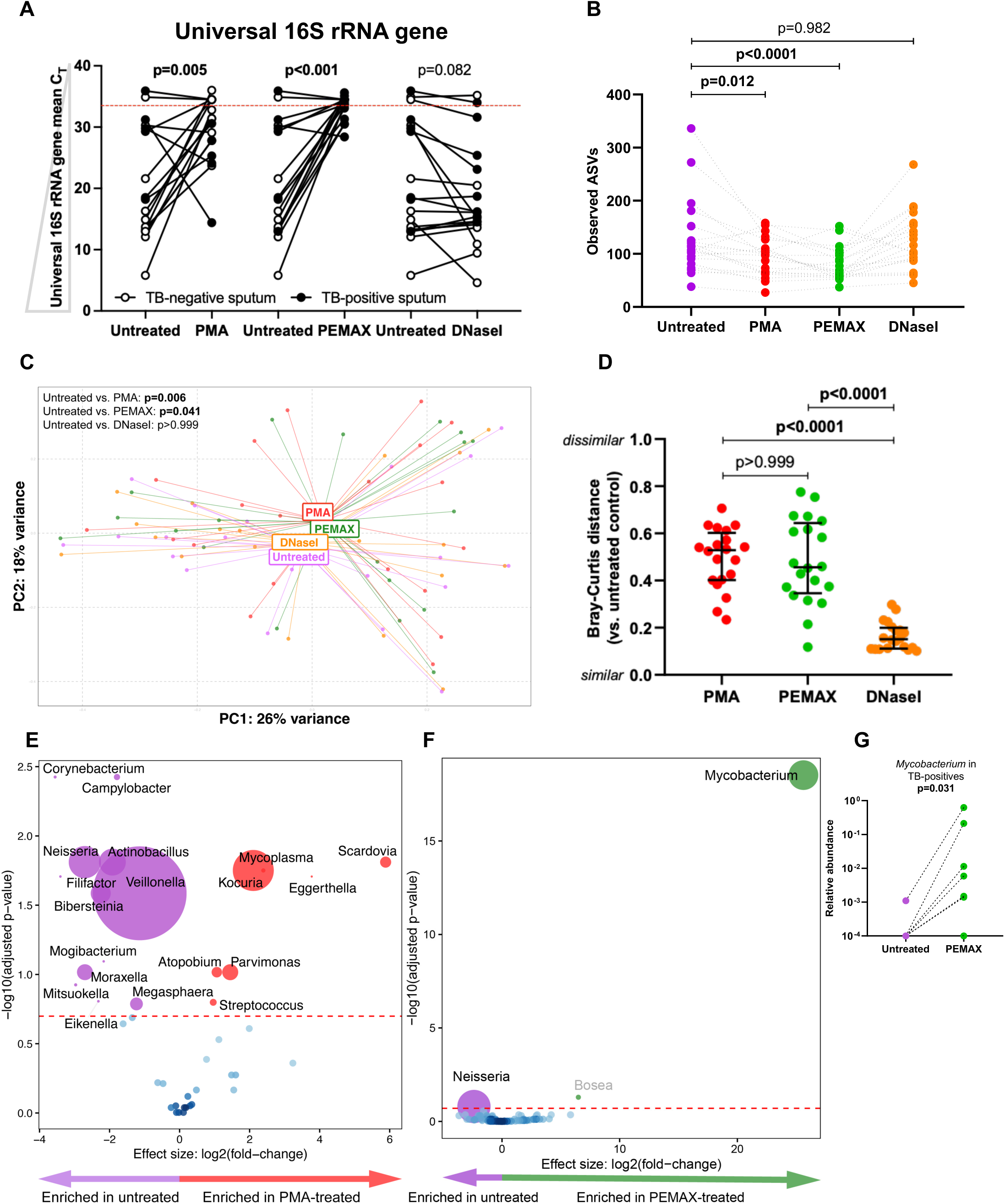
In sputa from people with presumptive TB, PMA or PEMAX reduces microbial diversity estimates more than DNaseI in comparison to untreated sputum. **(A)** PCR readouts for untreated versus treated (PMA, PEMAX, or DNaseI) decontaminated sputum from TB-negatives and TB-positives (Cohort A). Triangles on y-axes indicate decreasing load. **(B)** α-Diversity decreased after PMA and PEMAX treatment but not DNaseI. **(C)** β-diversity differences between untreated and treated groups. **(D)** Bray-Curtis distance to untreated sputum was higher in PMA- and PEMAX-treated sputum versus DNaseI-treated sputum. Volcano plots depicting **(E)** many differentially abundant taxa in PMA-treated versus untreated sputum and (F) *Mycobacterium*-enrichment and *Neisseria*-depletion in PEMAX-treated versus untreated sputa. More discriminatory taxa appear closer to the left or right, and higher above the threshold (red dotted line, false discovery rate of 0.2). Relative abundances correspond to circle size. **(G)** Use of PEMAX increased the proportion of *Mycobacterium* reads in TB-positives. ASVs: amplicon sequencing variants.

*Sequencing*: PMA- and PEMAX-treated sputum had lower α-diversity, whereas DNaseI-treated sputum had similar α-diversity to paired untreated controls (**Figure 3B**). β-diversity differed between untreated and PMA-treated (p=0.006), untreated and PEMAX-treated (p=0.041), and PMA-treated and DNaseI-treated sputum (p=0.019) (**Figure 3C**). Paired β-diversity distances between PMA- or PEMAX-treated and untreated sputum were greater than those between DNaseI-treated and untreated sputum (**Figure 3D**). Results remained similar when TB-positive and TB-negative groups were separately analysed (**Supplementary Figure 5A-B**).

#### PMA treatment reveals more taxa (versus untreated controls) than PEMAX, however, only PEMAX permits Mycobacterium detection

PMA permitted 19 more differentially abundant taxa to be identified vs its untreated controls (seven enriched, 12 depleted; *Scardovia* most enriched, *Corynebacterium* most depleted) (**Figure 3E**), than PEMAX [one enriched (*Mycobacterium*), one depleted (*Neisseria*), one identified as a potential contaminant (*Bosea*)] (**Figure 3F; Supplementary Figures 6-7**). None of the seven taxa enriched due to PMA were *Mycobacterium*. In PEMAX-treated sputum from people with TB, 6/9 (67%) had an increase in detected *Mycobacterium* relative abundance compared to untreated sputa (p=0.031) (**Figure 3G**) [5/9 (56%) of these would have not had any mycobacterial reads detected if PEMAX was not used]. No discriminatory taxa were associated with DNaseI treatment. After stratification by TB status, similar results with fewer differential taxa occurred **(Supplementary Figure 8)**.

#### PMA and PEMAX improve characterisation of microbiome differences by TB status compared to untreated controls

In untreated sputum, no α- and β-diversity differences occurred by TB status with *Gallibacterium* being the most discriminatory (enriched in TB-negatives). Upon PMA or PEMAX treatment, similar results occurred with *Scardovia* additionally enriched in TB-negatives (**Figure 4A-E**, **Supplementary Figure 9**). *Mycobacterium* was additionally enriched in TB-positives after PEMAX. A comparison of *Mycobacterium* relative abundances stratified by TB status and treatment method showed *Mycobacterium* enriched in PMA- and PEMAX-treated sputa from TB-positives (p=0.026 and p=0.002, respectively) (**Figure 4F**). No discriminatory taxa were found between patient groups with DNaseI treatment.

**Figure 4.**
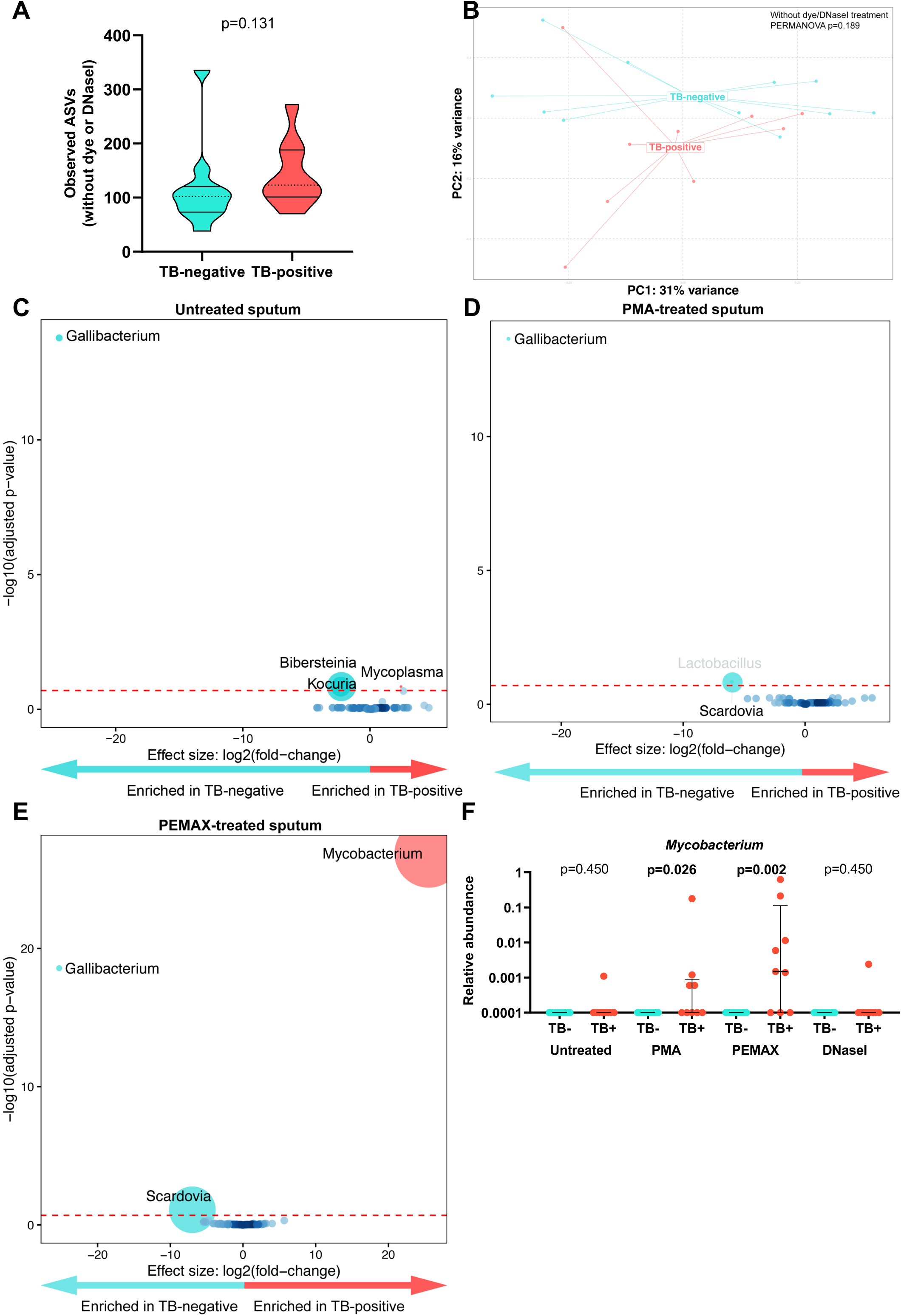
PMA and PEMAX enhances *Mycobacterium* detection in TB-positives versus TB-negatives. In untreated sputum, groups did not differ in **(A)** α-diversity nor **(B)** β-diversity. The same was observed for PMA-, PEMAX-, and DNaseI-treated sputum (see Supplement). TB-negatives were *Gallibacterium*-enriched in **(C)** untreated, **(D)** PMA-treated and **(E)** PEMAX-treated sputum versus TB-positives. More discriminatory taxa appear closer to the left or right, and higher above the threshold (red dotted line, false discovery rate of 0.2). Relative abundances correspond to circle size. **(F)** *Mycobacterium* relative abundance was higher in TB-positives than TB-negatives in sputum treated with either PMA or PEMAX, however, abundances were the same in untreated or DNase-treated sputum. In TB-positives, PEMAX permitted greater mycobacterial detection than PMA [0.0015 (0.0001-0.1125) vs. 0.0001(0.0001-0.0009)]. ASVs: amplicon sequencing variants.

### PEMAX treatment reveals diversity changes from TB chemotherapy otherwise undetected (Cohort B)

#### Comparisons within timepoints (untreated versus treated)

α-Diversity differed only at week 24, not week 0, between treated and untreated sputa, with lower α-diversity observed after PEMAX (**Figure 5A**; p=0.033). There were no β-diversity differences at week 0 nor week 24 (**Figure 5B**). No taxonomic differences were found at week 0 in treated versus untreated sputum, however, at week 24, treated sputa were *Neisseria*-enriched and *Mogibacterium*-depleted (**Figure 5C**).

**Figure 5.**
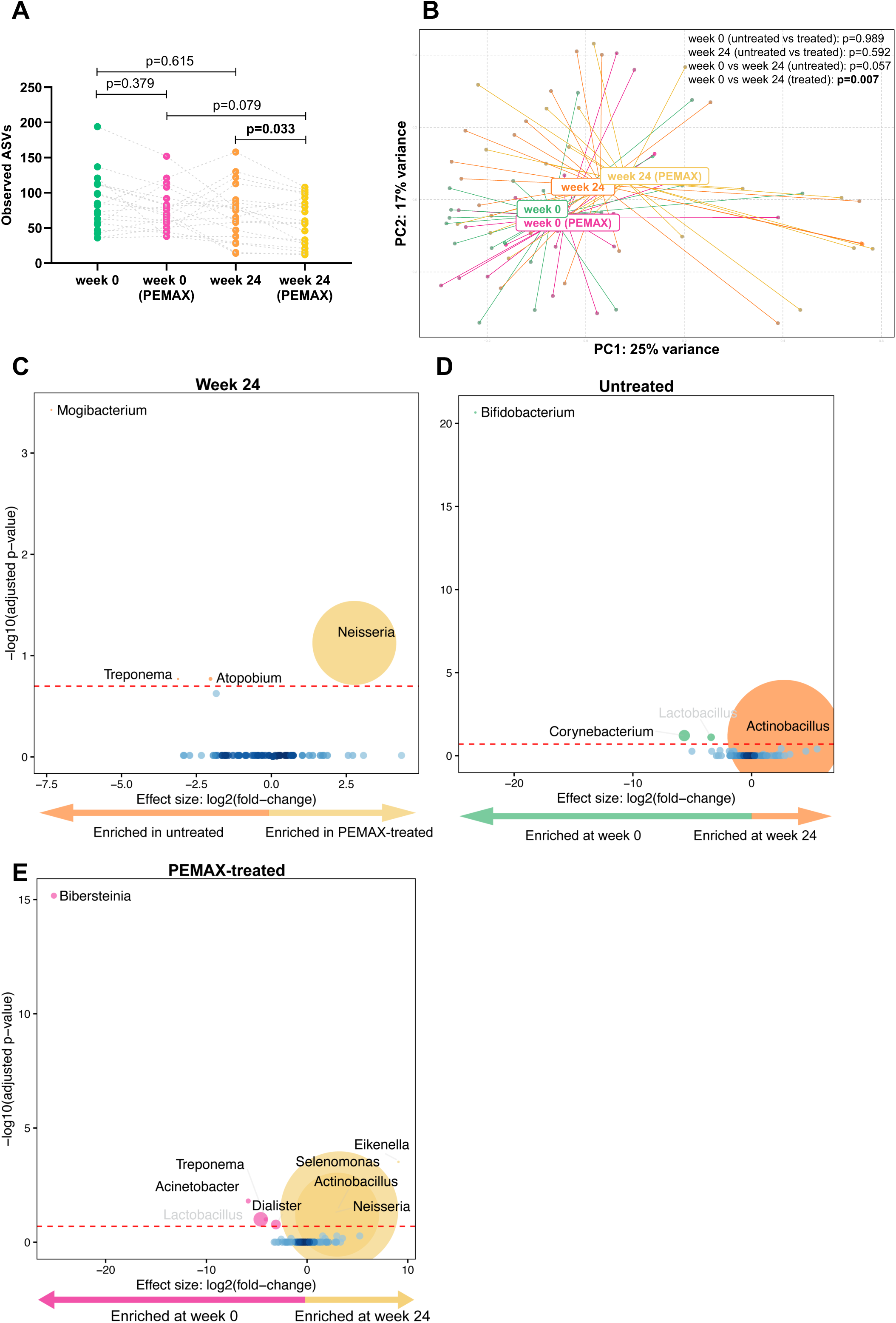
Microbial changes after TB chemotherapy are only detectable with PEMAX. **(A)** α-Diversity was only different between week 24 untreated and PEMAX-treated sputum. **(B)** Unlike in untreated sputum, PEMAX enabled detection of β-diversity changes between weeks 0 and 24. **(C)** *Neisseria*-enrichment and *Mogibacterium*-depletion at week 24 in treated vs. untreated sputum. **(D)** Only three discriminatory taxa were identified in the absence of PEMAX between weeks 0 and 24, whereas **(E)** many were identified after PEMAX-exposure. More discriminatory taxa are closer to the left or right, and higher above the threshold (red dotted line, false discovery rate of 0.2). Relative abundances of taxa are indicated by circle size. ASVs: amplicon sequencing variants.

#### Comparisons within treatment groups (week 0 versus week 24)

α-Diversity was unchanged per patient between timepoints (**Figure 5A**). β-diversity only changed over time in treated sputa (**Figure 5B**; p=0.007). In untreated sputa, *Bifidobacterium* and other taxa were enriched at week 0 vs. week 24 (**Figure 5D**). However, in treated sputum, these taxa were absent and other differences occurred (**Figure 5E**).

## Discussion

We demonstrated 1) PMA- and PEMAX-treated samples and sputum have lower detected mycobacterial load compared to untreated counterparts, 2) PMA- and PEMAX treatment of sputa reduces α-diversity and results in β-diversity differences compared to paired untreated sputum, 3) *Mycobacterium* is enriched in PEMAX-treated sputum from TB-positives compared to TB-negatives, a difference that was not seen in untreated sputum, 4) chemotherapy-associated microbiome changes are clearer (i.e., communities are compositionally dissimilar) in sputum treated with PEMAX versus those not treated, and 5) DNaseI treatment produces inconsistent signals in *in vitro* experiments and does not influence microbiome estimates. Together, these data have implications for characterising pathogen load and the microbiome in people with TB, including evaluating the effect of TB chemotherapy.

PMA and PEMAX improve differentiation of live from dead foodborne pathogens such as *Salmonella*, *Campylobacter*, and *Listeria*^42^. PMA also preferentially excludes dead rather than live *Mtb* in smear-positive sputa^27,29,43^, however, no studies have evaluated PEMAX in TB. Lower detected mycobacterial load was found with PMA and PEMAX treatment in PCR (qPCR, Xpert) implying some bacilli even from laboratory-generated dilution series are not intact. The effects of PMA, PEMAX, and DNaseI were not as strong as expected in the presence of increased cellular membrane disruption caused by antibiotics and heat killing (for example, PCR results becoming negative). This corroborates earlier Xpert data showing imperfect specificity and minimal improvement with PMA pre-treatment.^24^

Compared to paired untreated samples, PMA- and PEMAX-treated samples had lower microbial richness (i.e., suppressed DNA signals from non-intact bacteria). The inclusion of PMA or PEMAX is thus important for removing taxa likely not biologically active. These methods further identified *Mycobacterium* in people with TB that would otherwise go undetected. Previously, we demonstrated how a nested mycobacteriome approach could help to resolve this^8^; dye treatment represents an alternative. This is relevant for microbiome studies involving people with TB, as we and others have shown that *Mycobacterium* reads are often undetectable by 16S rRNA gene sequencing in sputum.^44,45^

Prolonged exposure to TB chemotherapy, particularly those with broader antimicrobial spectra that are common in treatment regimens for drug-resistant TB, and host immunological responses are likely to produce non-viable bacterial populations^12^. PEMAX improved detection of microbiome changes in sputum before and after chemotherapy by detecting more discriminatory taxa. It is worth considering if previously documented treatment-related sputum microbiome changes^46,47^ overlooked any taxonomic changes that could have been detected using a viability dye.

DNaseI performed poorly in dilution series and sputum microbiome evaluations. Prior research had suggested that the presence of antibiotics increased DNaseI sensitivity^48^, but this was not the case, which may be related to differences in specimen type or bacterial species.

The results must be viewed in the context of its limitations. Xpert assays was done using a dilution series of cells instead of sputum, in which cells are embedded within a matrix, which may affect results. Furthermore, sputum decontamination processes (commonly done as part of TB microbiology) can cause cell death while leaving cell membranes intact, in which case PMA and EMA have lower reduced ability to remove background DNA. This, however, is one reason we used PEMAX, which was designed to overcome this constraint^16^. Moreover, we used a modified sputum decontamination procedure^44^ in which sodium hydroxide was omitted to minimise effects on bacterial viability. Some study participants (Cohort B, who had MDR-TB) may have already been on chemotherapy at week 0, however, it is difficult to obtain a true baseline specimen from such people who are often prescribed first-line chemotherapeutics before drug resistance is discovered.

As sputum is an imperfect surrogate for the lower airways, many studies are shifting to bronchoalveolar lavage (BAL) sampling. Pre-treating low biomass specimens, such as BAL fluid or cough aerosols, where background signals from medical equipment or PCR reagents (or “reagent microbiome”) can interfere with biological interpretation,^51^ is likely to be beneficial, but further validation is required.

Our findings highlight the critical role of PMA or PEMAX pre-treatment in TB respiratory microbiome research, particularly during chemotherapy, as it provides invaluable insights into bacterial load characterisation and microbiome dynamics, especially when considering chemotherapeutics.

## Acknowledgements

This work was supported by the South African National Research Foundation (#98948) and the South African Medical Research Council (SAMRC) through its Division of Research Capacity Development under the SAMRC Intramural Postdoctoral programme from funding received from the South African National Treasury. The content hereof is the sole responsibility of the authors and do not necessarily represent the official views of the SAMRC or the funders. CN and research reported in this publication was supported by the Fogarty International Center of the National Institutes of Health under Award Number K43TW012302. The content is solely the responsibility of the authors and does not necessarily represent the official views of the National Institutes of Health. RV is supported by an NRF PDP Postdoctoral fellowship through the SAMRC, the DSI-NRF Centre of Excellence for Biomedical Tuberculosis Research and the Faculty of Medicine and Health Sciences at Stellenbosch University. GT acknowledges funding from National Institute of Allergy and Infectious Diseases of the National Institutes of Health under award number R01AI136894, EDCTP programme supported by the European Union (TMA-2015-SF-1041), and the SAMRC Intramural Flagship Project. Computations were performed using facilities provided by the University of Cape Town’s ICTS High Performance Computing team: https://hpc.uct.ac.za/.

## Data availability

The data that support the findings of this study will be openly available in the Sequence Read Archive at https://www.ncbi.nlm.nih.gov/sra. Other supporting data generated in this study are available from the corresponding author on reasonable request.

