## Supplement for "Subtracting the background: Reducing cell-free DNA’s confounding effects on *Mycobacterium tuberculosis* quantitation and the sputum microbiome"

[a]DSI-NRF Centre of Excellence for Biomedical Tuberculosis Research; South African Medical Research Council Centre for Tuberculosis Research; Division of Molecular Biology and Human Genetics, Faculty of Medicine and Health Sciences, Stellenbosch University, Cape Town, South Africa.
[b]African Microbiome Institute, Faculty of Medicine and Health Sciences, Stellenbosch University, Cape Town, South Africa.
[c]Municipal Laboratory – Aigües de Mataró, Mataró, Spain.
[d]Reactivos para Diagnóstico, Setmenat, Spain.
[e]Department of Psychiatry, Faculty of Medicine and Health Sciences, Stellenbosch University, Cape Town, South Africa.
[f]Institute of Infectious Disease and Molecular Medicine (IDM), Faculty of Health Sciences, University of Cape Town, Cape Town, South Africa.
[g]Department of Genetics and Genomic Sciences, Icahn School of Medicine at Mount Sinai, New York, NY, USA.
[h]Public Health Research Institute, New Jersey Medical School, Rutgers, The State University of New Jersey, Newark, NJ, USA.
[i]Division of Pulmonary, Critical Care, and Sleep Medicine, New York University School of Medicine, New York, USA.

[†]Contributed equally

**Supplementary Table 1. Primer sequences**

| Name | Sequence | Amplicon size (bp) |
|---|---|---|
| 16S universal | *515F* 5’-GTGCCAGCMGCCGCGGTAA-3’<br>*806R* 5’-GGACTACHVGGGTWTCTAAT-3’ | 292 |
| 16S MTB | *MtbF* 5’-GTGCCAGCAGCCGCGGTAA-3’<br>*MtbR* 5’-GGACTACCAGGGTATCTAAT-3’ | 292 |

**Supplementary Figure 1. Lower mycobacterial load readouts occurred in the antibiotic-exposed dilution series treated with PMA and PEMAX versus untreated controls.** This occurred for each CFU concentration tested (108, 106). Figure 2 has these data for the 24h exposure period. The non-template control is indicated by the red dotted line. Triangles on the y-axes indicate decreasing load. Data are median with IQR. MTBC: Mycobacterium tuberculosis complex; CT: cycle threshold; PMA: propidium monoazide; IQR: interquartile range.

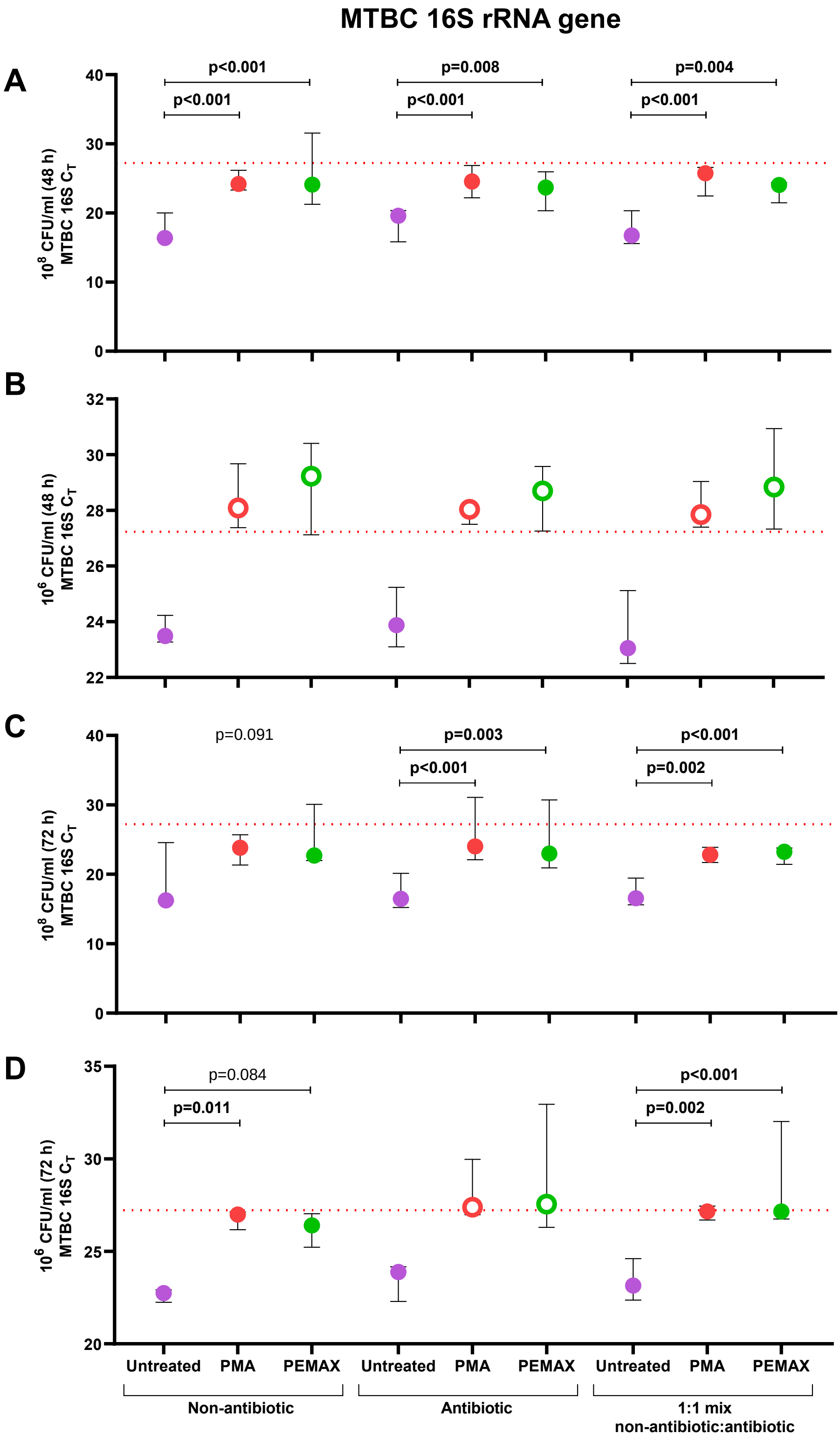

**Supplementary Figure 2. Lower mycobacterial load readouts occured after freeze-thaw in dilution series treated with PEMAX versus untreated controls.** This was tested for the 106 CFU/ml concentration from 24h, 48h and 72h incubation periods. The non-template control is indicated by the red dotted line. Triangles on the y-axes indicate decreasing load. Data are mean with 95% CI. CT: cycle threshold; FT: freeze-thaw. CI: confidence interval.

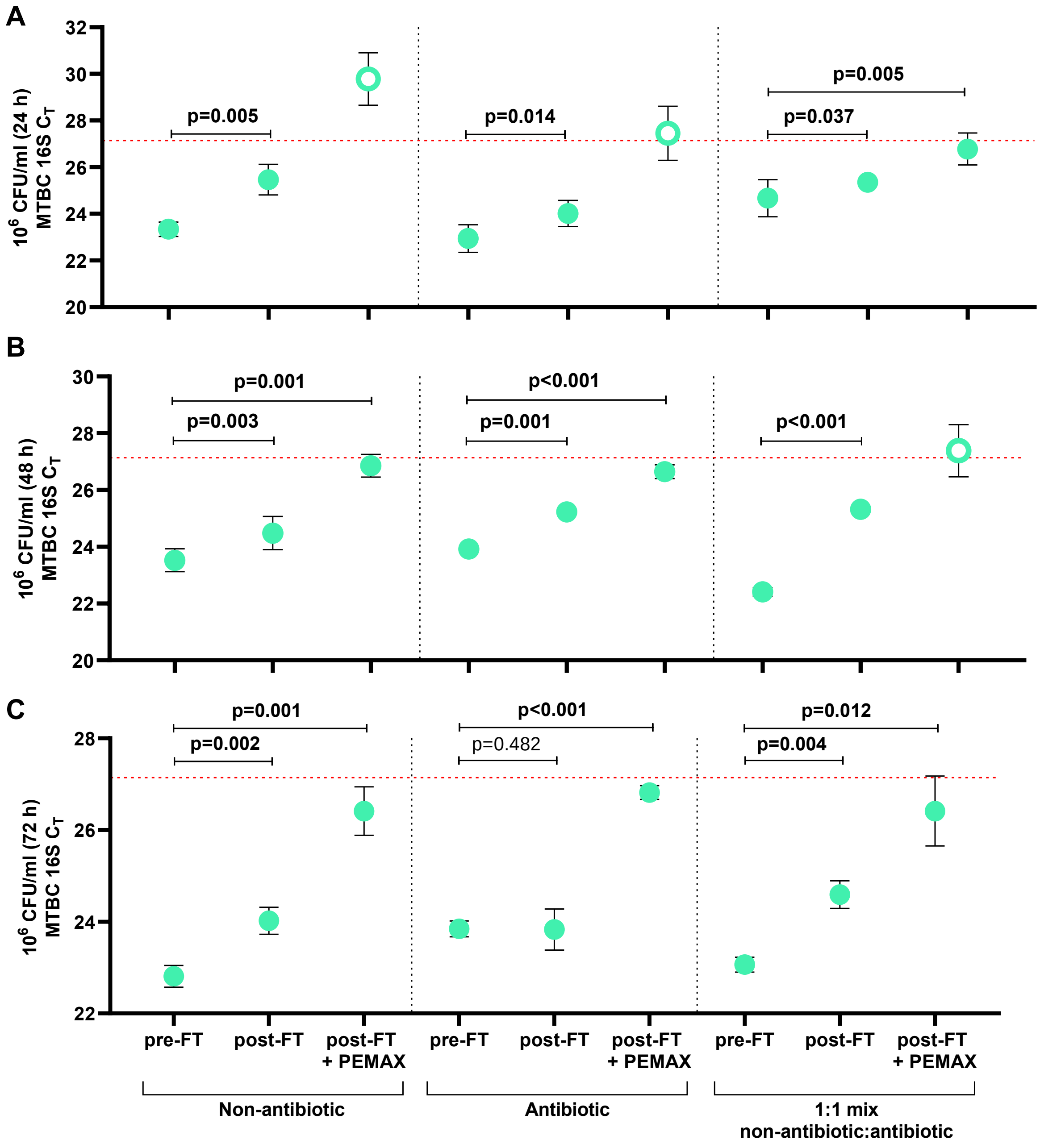

**Supplementary Figure 3. Similar mycobacterial load readouts in dilution series treated with DNaseI versus untreated controls.** This occurred for each CFU concentration tested (108, 106). The non-template control is indicated by the red dotted line for Experiment A and the grey dotted line for Experiment B. Triangles on the y-axes indicate decreasing load. MTBC: Mycobacterium tuberculosis complex; CT: cycle threshold.

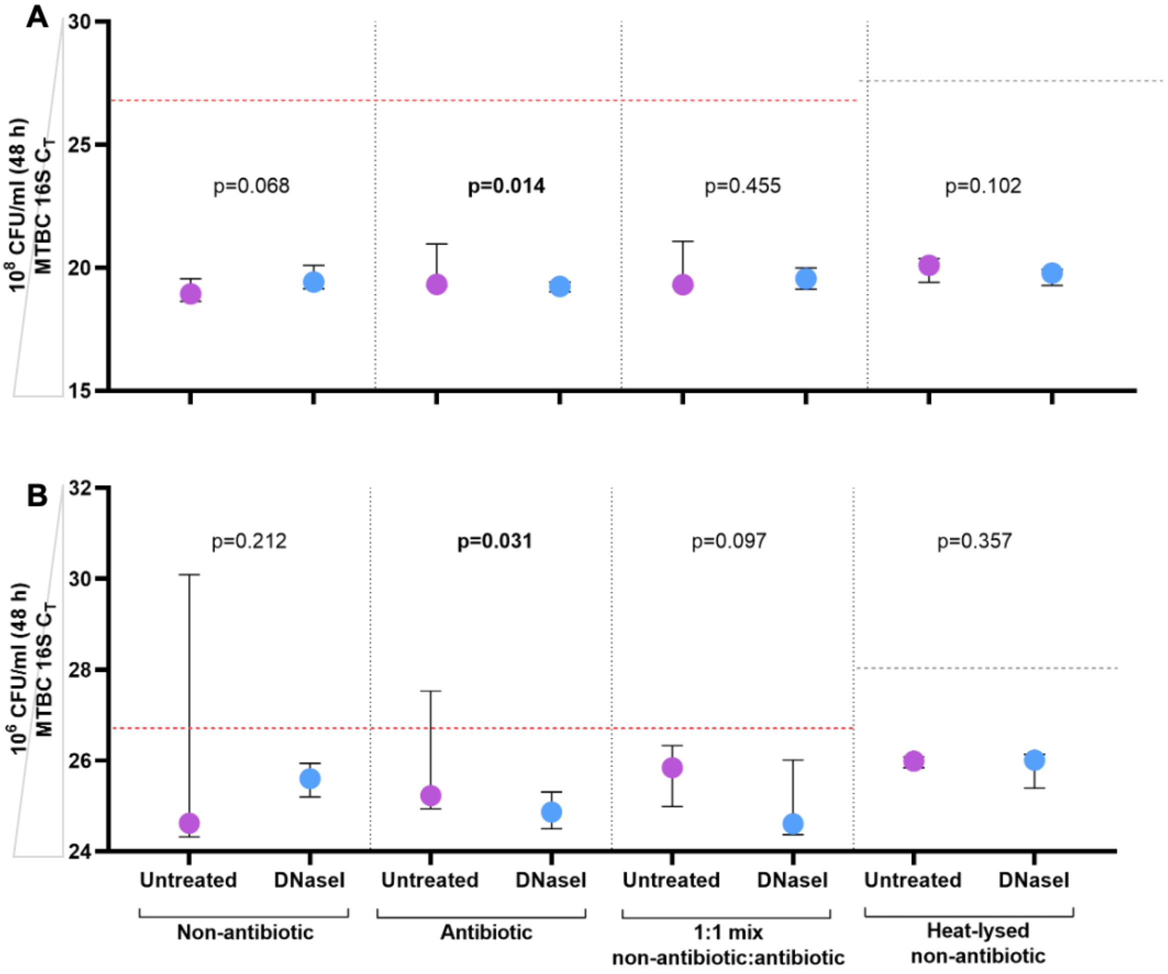

**Supplementary Figure 4. Similar mycobacterial load measured by Xpert MTB/RIF in dilution series treated with DNaseI versus untreated controls.** Median CTmin values are shown for different groups of diluted culture (104, 106, 108 CFU/ml) with vs. without DNaseI. CTmin: cycle threshold.

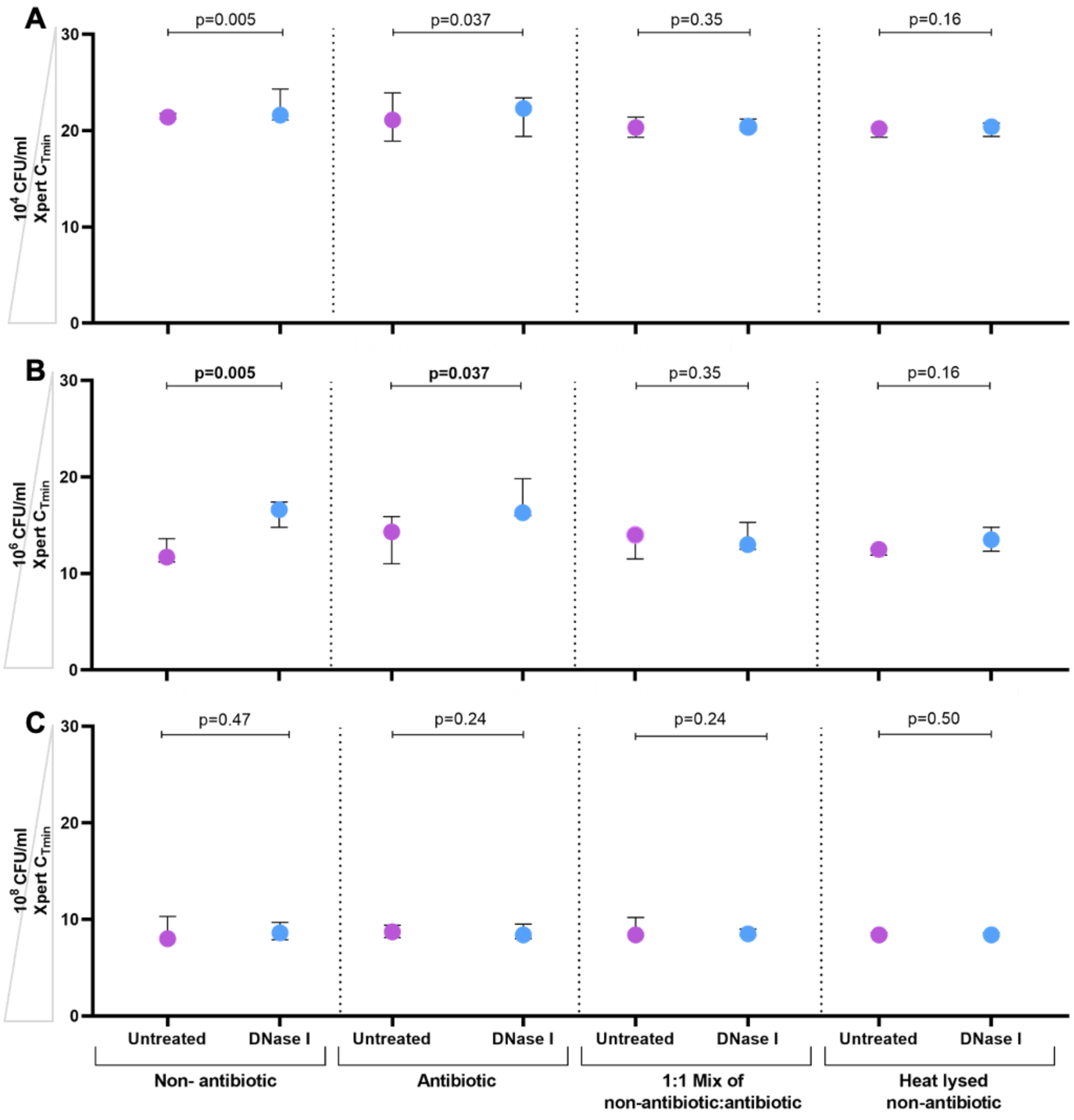

**Supplementary Figure 5. α- and β-diversity differences, stratified by TB status, between treated (PMA, PEMAX, DNaseI) and untreated sputum. Figure 3** has these data for all overall.

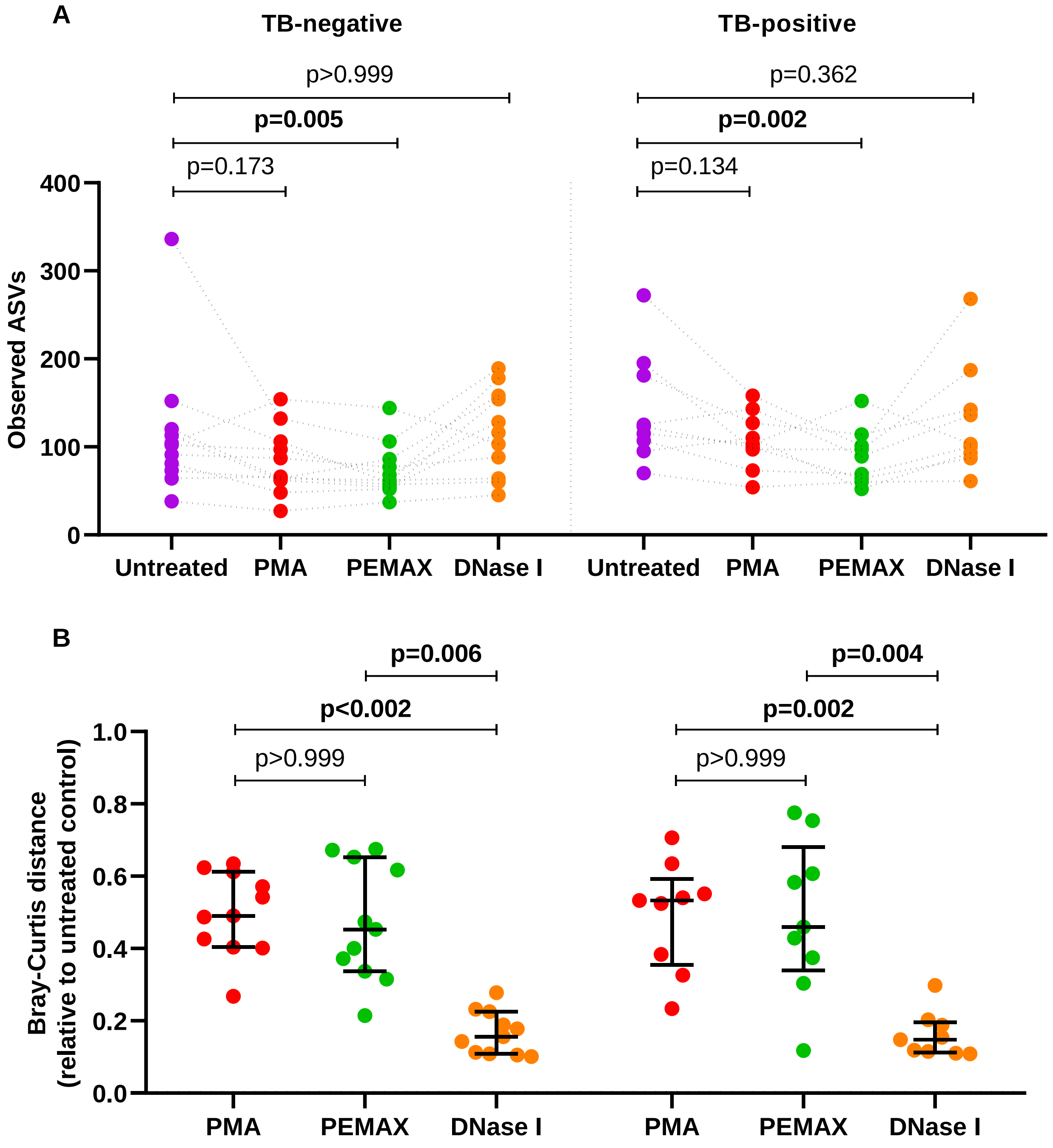

**Supplementary Figure 6. Potential contaminant ASVs in sputum vs. background based on the decontam prevalence method (ranked by sputum).** Mean relative abundances of ASVs in background and sputum, ranked by sputum (in decreasing order). ASV: amplicon sequence variant.

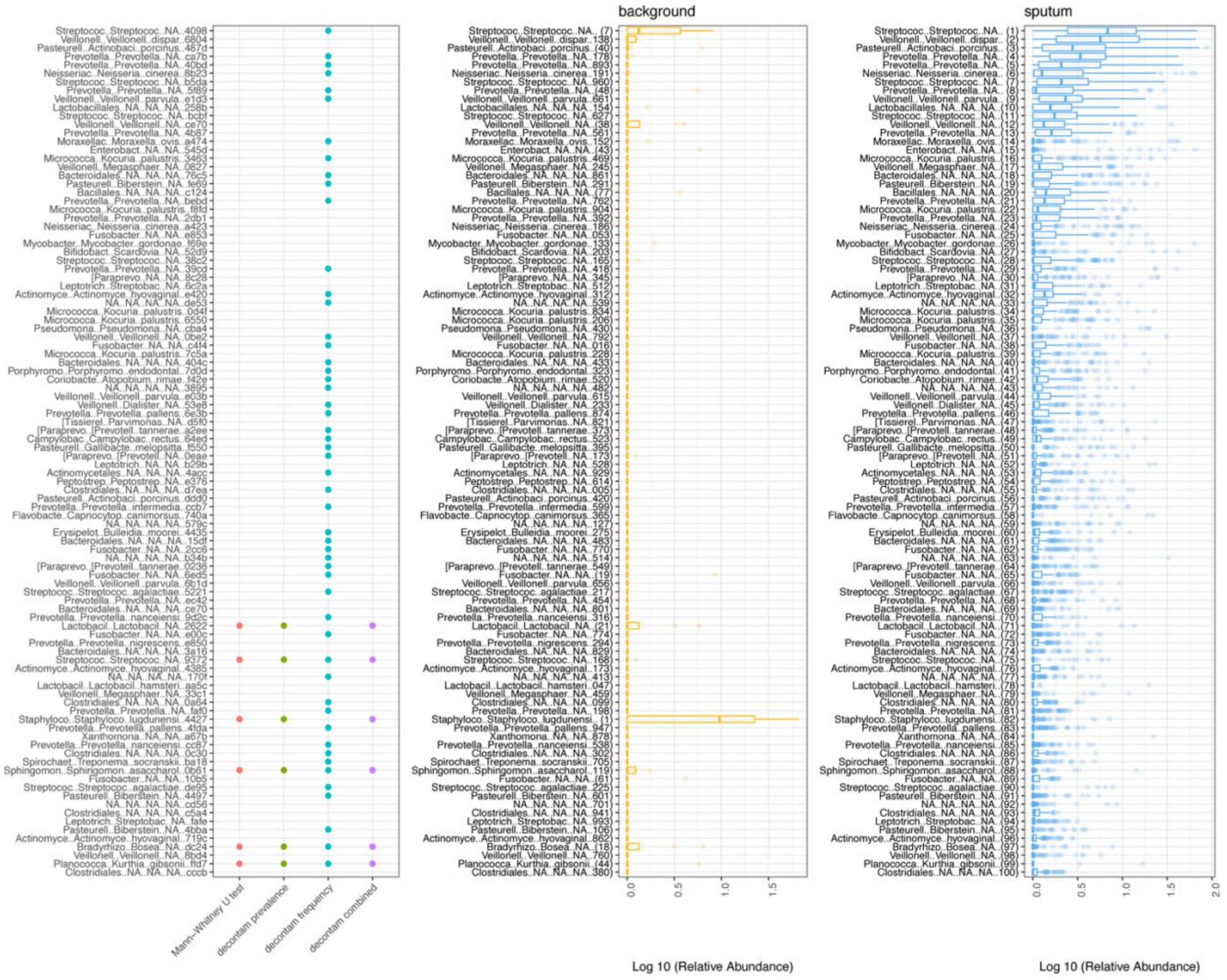

**Supplementary Figure 7. Potential contaminant ASVs in sputum vs. background based on the decontam prevalence method (ranked by background).** Mean relative abundances of ASVs in background and sputum, ranked by background (in decreasing order). ASV: amplicon sequence variant.

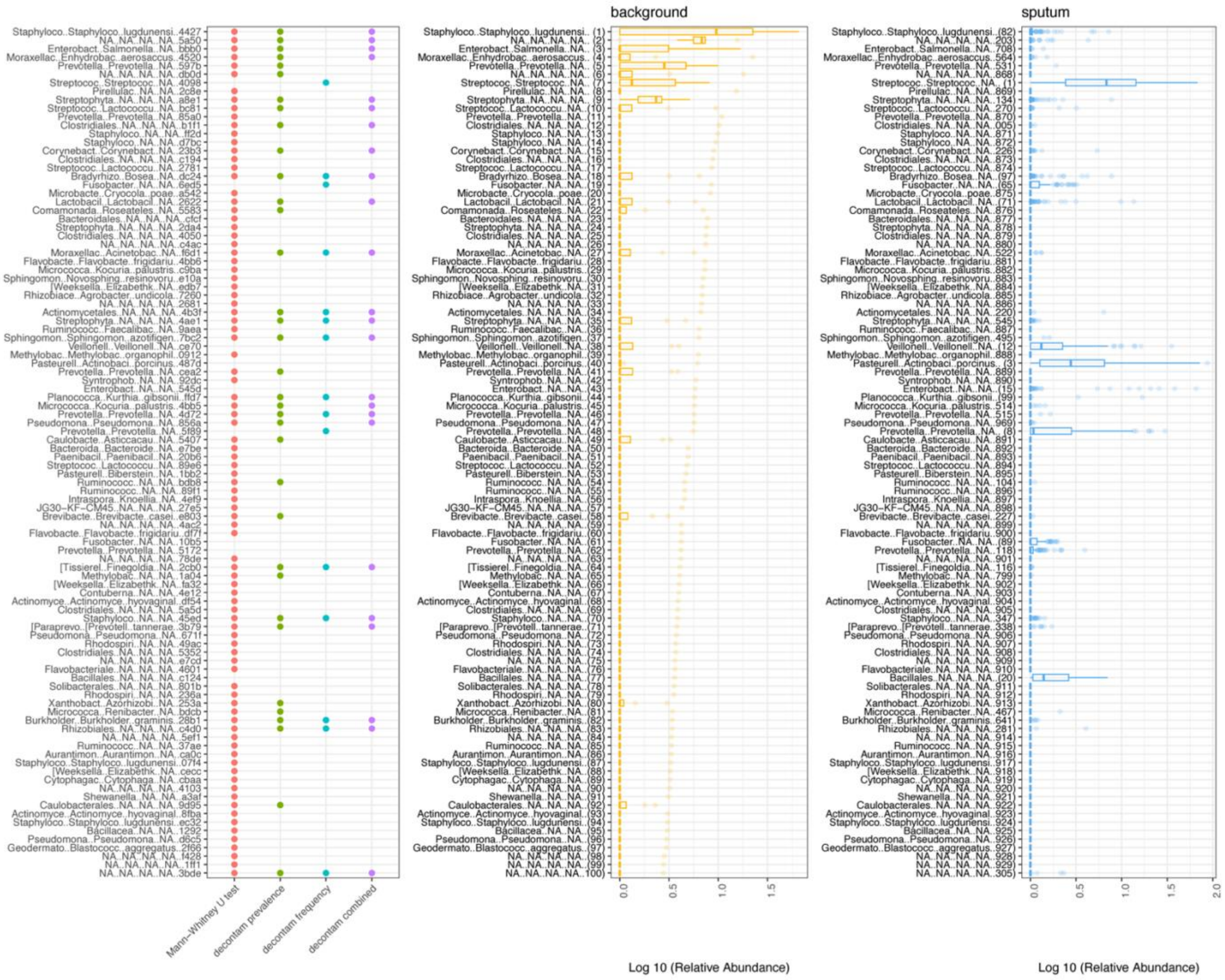

**Supplementary figure 8. Unlike PMA, PEMAX enables Mycobacterium detection in sputum from people with TB.** Volcano plots depicting discriminatory taxa identified after dye treatment. For PMA, (A) treated TB-negative sputum was *Scardovia*-enriched and *Corynebacterium*-depleted vs. untreated sputum (B) whereas treated TB-positive sputum was *Eggerthella*-enriched and *Actinobacillus*-depleted vs. untreated sputum. (C) For PEMAX, treated TB-positive sputum was *Mycobacterium*- and *Burkholderia*-enriched. More discriminatory taxa appear closer to the left or right, and higher above the threshold (red dotted line, false discovery rate of 0.2). Relative abundances correspond to circle size.

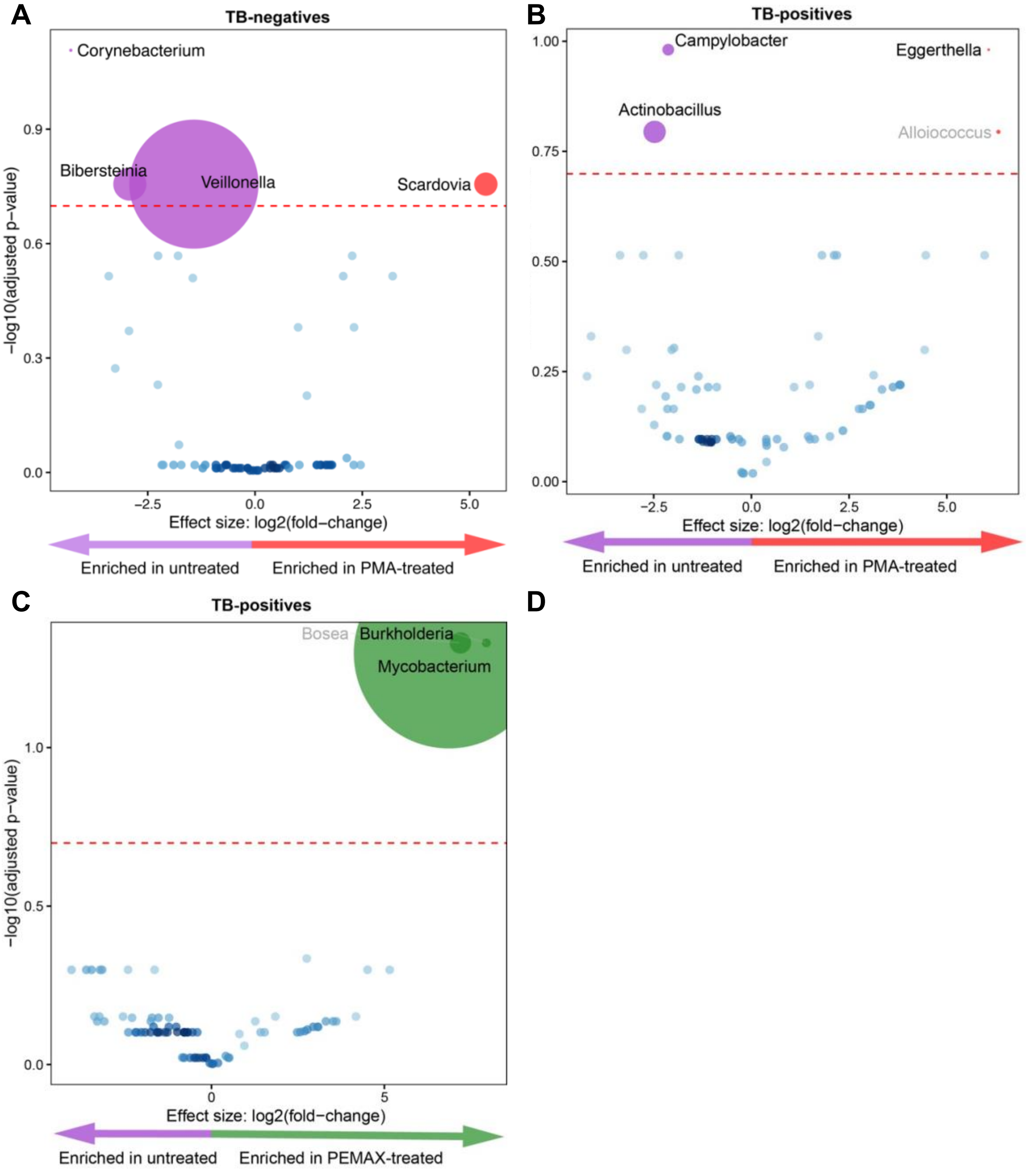

**Supplementary Figure 9. TB-negatives and TB-positives have similar α- and β-diversity irrespective of treatment (PMA, PEMAX, DNaseI). Figure 4** has these data for untreated sputum.

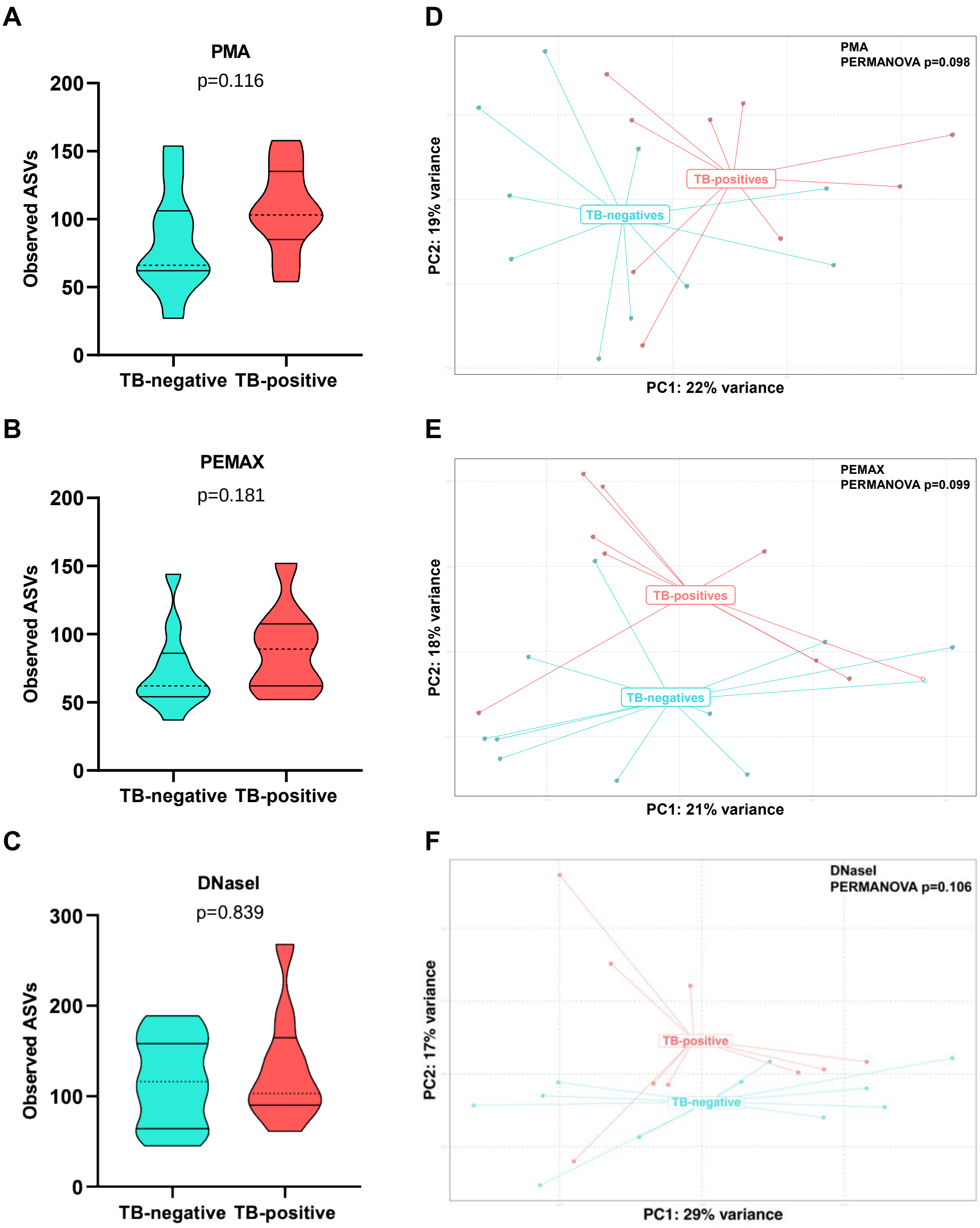
